# National diagnostic reference levels for digital diagnostic and screening mammography in Uganda

**DOI:** 10.1101/2023.11.05.23298120

**Authors:** Denish Odongo, Alen Musisi, Richard Omara Okello, Felix Bongomin, Geoffrey Erem

## Abstract

**Introduction:** Screening and diagnostic mammography are associated with some risk of radiation-induced breast cancer. This study was conducted to establish the National Diagnostic Reference Levels (NDRLs) for digital diagnostic and screening mammography in Uganda to achieve breast radiation dose optimization.

**Methods:** A cross-sectional study was conducted among female participants recruited by a consecutive sampling from three selected Hospitals with digital mammography in Uganda. The study variables extracted from the mammography machines were exposure factors, compressed breast thickness (CBT), and Average Glandular Dose (AGD) of two standard mammogram views. The stratified National DRL was derived by calculating 75^th^ percentile of the AGD across all the samples at various CBT ranges for both screening and diagnostic mammography in craniocaudal (CC) and mediolateral oblique (MLO) views.

**Results:** We included 300 participants with mean ages of 50.28±9.32 and 47.45±13.45 years for the screening and diagnostic mammography, respectively. There were statistically significant positive correlations between AGD and exposures factors (mAs, kVp) (all p-values<0.0001). For screening mammography, mAs demonstrated a strong positive correlation (r = 0.8369 in CC, 0.8133 in MLO), whereas kVp showed positive correlation with relatively lower coefficients (r = 0.3700 in CC, 0.3080 in MLO). In diagnostic mammography, mAs exhibited an even stronger positive correlation (r = 0.8987 in CC, 0.8762 in MLO), and kVp maintained a positive correlation with somewhat lower coefficients (r = 0.4954 in CC, 0.3597 in MLO). In screening mammography, for CBT within the range of (7-39)mm, the NDRLs were (1.5mGy, 1.66mGy) in CC) and MLO views. For CBT in the range of (40-59)mm, the NDRLs were (1.78mGy, 1.87mGy), and for CBT in the range of (60-99)mm, the NDRLs were (2.18mGy, 2.22mGy). For diagnostic mammography, the NDRLs were established as (1.7mGy, 1.91mGy), (2.00mGy, 2.09mGy), and (2.63mGy, 2.81mGy) for CBT ranges of (7-39)mm, (40-59)mm, and (60-99)mm, respectively.

**Conclusion:** The NDRLs for digital screening and diagnostic mammography in Uganda have been proposed for the first time. The NDRL values in mammography should be specific to CBT ranges and mammographic views for both diagnostic and screening mammography.

## Introduction

According to the World Health Organization (WHO), approximately 2.26 million new cases of breast cancer in women were diagnosed globally in 2020, accounting for one in every eight cancer patients making it the most prevalent cancer and is the leading cause of cancer death among women. However, this figure varies by regions, for example lung cancer was the leading cause of death in China though breast cancer was still the most prevalent cancer in that country in 2020 [1].

Modalities including ultrasound and MRI can be used for breast examinations but mammography is accepted to be the gold standard technique in breast cancer diagnostic studies [2]. The use of mammography has been shown to reduce deaths from breast cancer by 20-40% [3, 4]. Mammography is performed using either the screen film mammography (SFM) or digital mammography machine for screening in asymptomatic women at increased risk of breast cancer and diagnosis for women with symptoms suggestive of breast cancer [5].

In digital mammography, the x-rays transmitted through the breast are absorbed by an electronic detector, recorded and displayed using computer image-post processing as opposed to SFM [6].

One of the serious risks associated with mammography is radiation-induced breast cancers [7]. A low dose of irradiation causes DNA damage in mammary epithelial cells following repeated mammographic imaging [8, 9]. Breast tissue has a tissue weighting factor of 0.12 which makes it one of the most radiosensitive tissues in the body with increased risk of the stochastic effect of radiation (**ICRP, 2007**).

The probability of occurrence of radiation-induced breast cancer increases with increasing average glandular dose (AGD) [10].

In 1996, the International Commission on Radiological Protection (ICRP) introduced the Diagnostic Reference Level (DRL) to optimize the dose for all ionizing radiation including mammography [11]. The DRL was purposefully introduced to help control the high radiation dose which increases the risk of radiation-induced breast cancer in women [12]. DRL is an optimum range of doses that is safer for patients to undergo examination while not losing out the diagnostic value of the image and it must be established for each imaging modality to help detect the unusual level of doses given to patients [13]. Glandular dose (GD) is appropriate to use to establish DRL quantity in mammography even though it is the measure of the organ dose rather than the amount of ionizing radiation used to perform the mammography. The DRL for mammography is generally derived from calculating the 75^th^ percentile of the distribution of AGD measurements from the observed sample.

There is no established national or local DRL for digital mammography in Uganda despite 18 mammography machines currently registered with the Atomic Energy Council. We aimed to establish the national DRLs (NDRLs) for both screening and diagnostic mammography in Uganda as well as establish the relationship between the AGD and the radiation exposure factors.

## Material and method

### Study design

This was a cross-sectional study design conducted for 2 years from the 1^st^ of January 2021 to the 31^st^ of December 2022.

### Study setting

Purposive sampling method was used to select three (60%) of the 5 digital mammography machines in Uganda. The ICR P 135 requires 35-50% for the initial establishment of the national DRL [11]. These hospitals were kept anonymous for confidentiality purposes by assigning the codes A, B and C. Two of these hospitals were located in Kampala and one was located in Jinja City. There was no functional digital mammography machine in the government hospital during this study period.

### Sample size estimation

According to the ICRP 135 of 2017 updated in 2019, data on DRL quantities in mammography requires a minimum of 50 patients per mammography machine [11]. Data for 50 participants were collected for each of the 3 study Hospitals per indication (screening and diagnostic mammography). The total number of participants for this study was (50x3x2) =300. For each participant, 4 breast mammographic views (RMLO, LMLO, RCC, and LCC) were considered. Therefore, the total mammographic views (images) for this study was (4x300) =1200.

### Sampling method

A consecutive sampling method was used to select records of 50 females who underwent screening and 50 female for diagnostic mammography from hospitals **A, B** and **C** and a total of 300 participants were selected.

### Data tool and data collection

The information about the machine model, manufacturers, year of manufacturing and year of installation was recorded in the data abstraction form. The study variables for the two views mammographic images of the left craniocaudal (LCC), right craniocaudal (RCC), left mediolateral oblique (LMLO), and right mediolateral oblique (RMLO) were retrospectively accessed from 25^th^ April-2023 to 25^th^ May-2023 and extracted from the mammography Digital Imaging and Communications in Medicine (DICOM) header then recorded in a data abstraction form. The Image quality was graded using PGMI (Perfect, Good, Moderate and Inadequate) method for evaluation of the clinical image quality in mammography [14]. The most qualified and experienced radiographers from every centre were assigned to evaluate the image quality categorized them into PGMI [15] and recorded them in a data collection tool.

### Study variables

The age of patients, kilovoltage peak (kVp), milliampere-seconds (mAs), compressed breast thickness (CBT) in millimetres (mm), anode target/filter combinations, entrance surface dose (ESD) in milli Gray (mGy) and AGD in mGy. In this study, the Average Glandular Dose (AGD) for each acquired image was calculated automatically in the mammographic machine and recorded in the system using the methods described by Dance et al. [16–18].

### Data Management and Analysis

The data from the data collection tool were entered manually into the Excel sheet. The AGD, CBT, BCF, ESD, mAs and kVp of the right craniocaudal (RCC) and left craniocaudal (LCC) views of the breast images were summed up and divided by two to obtain their mean for the craniocaudal (CC) view. A similar process was repeated for the mediolateral oblique (MLO) view. These were then exported to STATA version 17 software and analyzed. Descriptive statistics were used to obtain the mean, median, standard deviation, percentiles, and range values. AGD relationship with the exposure factors (kVp, mAs), ESD, CBT and Breast Compression Force (BCF) was tested using a Pearson correlation coefficient test and presented in the table form. The values of the CBT were grouped into three categories of a narrow range which were representative of CBT for the Ugandan women population as recommended by ICRP number 135 for every country to determine their women’s CBT ranges [11, 19].

The NDRL values of the screening and diagnostic mammography were determined by calculating the 75^th^ percentile of the median values of the AGD across the identified CBT ranges in each mammographic view (CC and MLO) from the combined data across all the 3 hospitals. A comparison of established NDRL in these study with the NDRLs of other countries was made and presented in a table form.

### Quality Assurance and quality control

Quality control tests were performed by a qualified medical physicist and radiographer for all the 3 mammographic machines. These tests were daily digital tests with monitor checks, weekly digital tests requiring homogeneity (image quality) checks, weekly checks of automatic exposure control and 3-6 monthly tests, performed by a medical physicist which include kVp accuracy and reproducibility, output linearity etc.

The research assistants were adequately trained and routinely supervised by the principal investigator to ensure the correct use of the data collection tool and adherence to ethical principles. The completed abstraction forms were checked and verified with the data from the machine for completeness and accuracy.

### Ethical considerations

Ethical approval to conduct the study was obtained from the Institutional Review Board (IRB) of the School of Medicine, Makerere University College of health sciences (protocol reference number: Mak-SOMREC-2023-562). The IRB also provided a waiver of consent by the participant since it was a retrospective data collection. Permission to access patients’ data and their mammographic images were obtained from the executive directors of the respective hospitals which provide mammographic services. All the information and mammogram were kept confidential and used only for the present study by keeping all study materials under lock and key. Anonymity was maintained for both the participating hospitals and study participants by using alphabetical letters and serial numbers for hospitals and patients other than their names respectively.

## Results

In this study, three (3) out of the five (5) digital mammography machines were selected from three Hospitals A, B and C, accounting for 60% of all digital mammography machines in Uganda.

### Results on mammography machines for hospitals A, B and C

Hospitals A and C had Senographe digital mammography machines with similar voltage (220-230V) and frequencies (50/60Hz). Their power inputs were 6.9kVA and 7kVA respectively.

Hospital B had a Siemens digital mammography. Its voltage is 220-230V, frequency of 50/60Hz, and power input of 7.5kVA. All hospitals in this study used a large focal spot size (0.3mm) and tungsten/Rhodium (W/Rh) anode target/filter combination to acquire mammography images for both screening and diagnostic mammography (Table 1).

**Table 1.** The Digital mammography machine from Hospitals A, B and C.

| **Mammography**<br>**Machine parameters** | **Hospitals** |  |  |
| --- | --- | --- | --- |
|  | **A** | **B** | **C** |
| **Ownership** | Private | PNFP | Private |
| **Type** | DR | DR | DR |
| **Model/make** | Senographe Crystal<br>Nova | Mammomat<br>inspiration Prime | Senographe, Crystal<br>Nova |
| **Manufacturer** | GE Medical System,<br>China | Siemens Health Care,<br>Germany | GE Medical System<br>Korea |
| **Year of manufacture** | June/2020 | October/2019 | December/2018 |
| **Year of installation** | 2021 | 2020 | 2020 |
| **Focal spot size used** | 0.3 mm | 0.3 mm | 0.3 mm |
| **Anode/filter**<br>**combination used** | Tungsten/Rhodam<br>(W/Rh) | Tungsten/Rhodam<br>(W/Rh) | Tungsten/Rhodam<br>(W/Rh) |

Where, **DR** = Digital Radiography, **GE**=General Electric; **PNFP** – private not for profit

### Demography of the study participants

In Table 2, the age range of the participants in screening mammography was 33-76 years with a mean age of 50.28 ± 9.32 SD. While in the diagnostic mammography, the age range was 15-84 years with a mean age of 47.45 ±13.45 SD.

**Table 2.** The number of participants under 30 years, 30-44 years and ≥45 years of age (N=300)

| Age (years) | Screening Mammography<br>(n=150) | Diagnostic Mammography<br>(n=150) |
| --- | --- | --- |
| <30 | 0 (0%) | 6 (4%) |
| 30 to 44 | 44(29.33%) | 64 (42.67%) |
| $\geq 45$ | 106 (70.67%) | 80(53.33%) |
| **Total** | **150** | **150** |
| **The mean, standard deviation and ranges of the age of the participants** |  |  |
| Mean±SD | 50.28±9.32 | 47.45±13.45 |
| Range | 33-76 | 15-84 |

Source: Data from hospitals A, B and C (N=150 Screening and 150 diagnostics, total=300).

### The relationship of AGD with the kVp, mAs, CBT, BCF and ESD

The relationship of the AGD with mammographic radiation exposure factors (kVp and mAs), CBT, BCF and Entrance Surface Dose (ESD) was assessed using the Pearson correlation coefficient test (Table 3).

**Table 3:** Pearson correlation coefficient for the relationship between the AGD with kVp, mAs, ESD, CBT, and BCF.

| Modality | Variables | Projections | Pearson correlation coefficient (r) | 95% CI | p-value |
| --- | --- | --- | --- | --- | --- |
| Screening | $\bar{K}Vp$ | CC | 0.3700 | 0.223, 0.501 | <0.0001* |
|  |  | MLO | 0.3080 | 0.155, 0.446 | <0.0001* |
|  | mAs | CC | 0.8369 | 0.781, 0.879 | <0.0001* |
|  |  | MLO | 0.8133 | 0.751, 0.861 | <0.0001* |
|  | ESD | CC | 0.8860 | 0.846, 0.916 | <0.0001* |
|  |  | MLO | 0.6689 | 0.570, 0.749 | <0.0001* |
|  | CBT | CC | 0.3783 | 0.232, 0.508 | <0.0001* |
|  |  | MLO | 0.4437 | 0.305, 0.564 | <0.0001* |
|  | BCF | CC | -0.1993 | -0.348, -0.040 | 0.0145* |
|  |  | MLO | -0.0935 | -0.250, 0.068 | 0.2552 |
| Diagnostic mammography | kVp | CC | 0.4954 | 0.364, 0.607 | <0.0001* |
|  |  | MLO | 0.3597 | 0.212, 0.492 | <0.0001* |
|  | mAs | CC | 0.8987 | 0.863, 0.926 | <0.0001* |
|  |  | MLO | 0.8762 | 0.833, 0.909 | <0.0001* |
|  | ESD | CC | 0.9168 | 0.887, 0.939) | <0.0001* |
|  |  | MLO | 0.9063 | 0.873, 0.931 | <0.0001* |
|  | CBT | CC | 0.3932 | 0.249, 0.521 | <0.0001* |
|  |  | MLO | 0.3857 | 0.240, 0.514 | <0.0001* |
|  | BCF | CC | 0.0598 | -0.101, 0.218 | 0.4676 |
|  |  | MLO | 0.0220 | -0.139, 0.182 | 0.7894 |

There was a strong positive correlation between AGD and mAs in both views of the screening mammography with r =0.8369 and 0.8133 (all p-values <0.0001) in both CC and MLO projections. For diagnostic mammography, the r=0.8987 and 0.8762 (allp-values <0.0001) in both CC and MLO projections. A strong positive correlation was seen with ESD too. The rest of the exposure factors showed a moderate positive correlation with the AGD for both modalities and mammographic views except for BCF which showed a negative correlation with the AGD in the CC mammographic view in the screening mammography (r = -0.1993, p = 0.0145).

### Description of the digital Mammographic variables among hospitals

In Table 4, the mAs range and means for hospital B were comparable to those of hospital A in both screening and diagnostic mammography and views. The mAs range and mean values for Hospital C were higher than hospitals A and B in the CC and MLO views for both screening and diagnostic mammography. Hospital C had the least BCF. The ranges of BCF (N) in the CC view were 30-150N for both the screening and diagnostic mammography as well as diagnostic MLO and it was 35-150N in diagnostic CC. The ESD mean, median, minimum and maximum ranges in this hospital C were higher than those in hospitals A, and B.

**Table 4.** Mean, Median, SD, range of mAS, BCF and ESD for Hospital A, B, C and ALL.

| Hos-<br>pital | Views | Modality | mAs |  | BCF (N) |  | ESD (mGy) |  |
| --- | --- | --- | --- | --- | --- | --- | --- | --- |
| | | | Mean ±SD | Range | Mean±SD | Range | Mean±SD | Range |
| **A** | CC | Screening | 91.84 ±30.07 | 36.95-134. | 98.1±33.65 | 30-170 | 4.68±1.9 | 1.2-8.69 |
| | | Diagnostic | 115.0 1±43.03 | 30.15-218 | 133.7±48.1 | 45-200 | 6.22±3.0 | 1.05-13.49 |
| | MLO | Screening | 105.88 ±39.85 | 37.1-221 | 114±40.18 | 50-200 | 6.21±4.9 | 0.96-6.24 |
| | | Diagnostic | 126.73 ±46.83 | 36.8-250 | 145.9±41.2 | 35-200 | 7.03±3.4 | 1.37-15.11 |
| **B** | CC | Screening | 83.09±24.61 | 37.2-130.05 | 97.41±14.8 | 58-133.5 | 3.84±2.1 | 1.25-11.95 |
| | | Diagnostic | 91.24.01±38 | 40.25-212.65 | 95.58±9.58 | 79-135.5 | 3.55±1.4 | 0.85-6.55 |
| | MLO | Screening | 103.59±37.78 | 27.85-206.4 | 106.47±22 | 77-165 | 4.75±2.0 | 0.7-10.8 |
| | | Diagnostic | 109.97±31.61 | 43.75-172.55 | 100.33±18 | 72.5-171 | 5.17±2.5 | 1.35-15.95 |
| **C** | CC | Screening | 129.35±36.16 | 73.8-155.7 | 78.8±30.04 | 30-150 | 6.22±2.0 | 2.24-8.49 |
| | | Diagnostic | 91.24.01±38 | 40.25-212.65 | 78±39.45 | 30-150 | 6.62±2.3 | 3.02-13.61 |
| | MLO | Screening | 143.30±46.24 | 60.5-192.5 | 88.1±31.83 | 35-150 | 6.92±2.7 | 2.56-13.85 |
| | | Diagnostic | 109.97±31.61 | 43.75-172.55 | 84.02±39.7 | 30-150 | 7.68±3.0 | 3.04-15.26 |
| **ALL** | CC | Screening | 98.29±34.26 | 36.95-184 | 91.44±28.68 | 30-170 | 4.81±2.1 | 0.85-10.72 |
| | | Diagnostic | 111.86±43.09 | 30.15-252.9 | 102.43±43.0 | 30-200 | 5.56±2.8 | 1.05-13.49 |
| | MLO | Screening | 113.43±40.92 | 27.85-222.5 | 102.86±33.8 | 35-200 | 5.96±3.5 | 0.7-6.4 |
| | | Diagnostic | 26.67±44.04 | 36.8-253 | 110.08±43.3 | 30-200 | 6.63±3.1 | 1.35-16.48 |

Meanwhile, the overall mean ESD values were lower in CC and higher in MLO for both screening and diagnostic mammography. This trend was not seen in the overall mAs and BCF.

### The National Diagnostic Reference Levels (DRLs) and kVp at various CBT ranges for the Screening mammography

In Table 5, the national DRL values at stratified CBT ranges per mammographic views for digital screening mammography were computed as the 75^th^ percentile/3^rd^ quartile of the median values of the AGD of all samples across all the hospitals. The national DRLs at CBT ranges of (7-39) mm, (40-59) mm and (60-99) mm were (1.5, 1.66) mGy, (1.78, 1.87) mGy, and (2.18, 2.22) mGy in CC and MLO views respectively. In the same table 4, the mean CBT was higher in MLO than in CC view for all the categories of ranges. The mean kVp increased with CBT and was higher in MLO than in CC projections. The ranges of AGD were (0.45-3.27) mGy.

**Table 5.** The mean, standard deviation, and range of kVp and AGD plus the 75^th^ percentile of median values of AGD at various CBT ranges for all participants.

| Modality | CBT Range | View | NO_ of images | CBT (mm) | kVp |  | AGD |  |  |  |
| --- | --- | --- | --- | --- | --- | --- | --- | --- | --- | --- |
|  |  |  |  | Mean±SD | Mean±SD | Range | Mean ±SD | Media n | Range | 75<sup>th</sup> percentile (NDRL) |
| **Screening** | 7-39 | CC | 23 | 28.34±8 | 27.41 ±0.69 | 26-29 | 1.23±0.40 | 1.28 | 0.51-2.01 | **1.54** |
|  |  | MLO | 19 | 28.95±7 | 27.62 ±0.70 | 25.5-28 | 1.32±0.43 | 1.31 | 0.45-1.89 | **1.66** |
|  | 40-59 | CC | 91 | 49.49±5 | 28.64 ±0.46 | 27.5-29 | 1.44±0.52 | 1.51 | 0.53-2.81 | **1.78** |
|  |  | MLO | 70 | 50.59±4 | 28.69 ±0.46 | 27.5-30 | 1.54±0.50 | 1.58 | 0.72-2.7 | **1.87** |
|  | 60-99 | CC | 36 | 66.31±4 | 29.74 ±1.04 | 29-34.5 | 1.76±0.58 | 1.87 | 0.98-2.74 | **2.18** |
|  |  | MLO | 61 | 69.52±8 | 29.82 ±0.70 | 28.5-32 | 1.85±0.66 | 1.78 | 0.6-3.27 | **2.22** |
| **Diagnostic** | 7-39 | CC | 23 | 31.43±7 | 27.67 ±0.76 | 26-29 | 1.38±0.53 | 1.43 | 0.55-2.64 | **1.7** |
|  |  | MLO | 16 | 31.31±7 | 27.88±0.85 | 26.5-29 | 1.52±0.70 | 1.45 | 0.59-3.05 | **1.91** |
|  | 40-59 | CC | 90 | 49.57±5 | 28.66±0.48 | 27.5-29 | 1.61±0.64 | 1.59 | 0.73-3.23 | **2.00** |
|  |  | MLO | 78 | 51.19±5 | 28.82±0.53 | 27.5-30 | 1.69±0.62 | 1.68 | 0.68-3.47 | **2.09** |
|  | 60-99 | CC | 37 | 66.49±5. | 29.61±0.49 | 29-31 | 2.04±0.74 | 2.13 | 0.95-3.55 | **2.63** |
|  |  | MLO | 56 | 70.04±7 | 29.96±0.77 | 29-32 | 2.16±0.74 | 2.22 | 1.09-3.81 | **2.81** |

\*p<0.05

### The National Diagnostic Reference Levels and kVp at various CBT ranges for the Diagnostic mammography

Similar to that of screening mammography, the NDRLs for diagnostic mammography at CBT ranges of (7-39) mm, (40-59) mm and (60-99) mm were (1.7, 1.91) mGy, (2.00, 2.09) mGy, and (2.63, 2.81) mGy respectively. The national DRL values for diagnostic mammography were also higher than those for screening mammography for the corresponding CBT and views.

The trend of the CBT and kVp were similar but had higher values than those in the screening mammography of the corresponding CBT and views (Table 5).

### Image quality assessment

The **PGMI** (Perfect, Good, Moderate and Inadequate) method for evaluation of the clinical image quality in mammography [14] was used. The overall result indicated that more than 50% of all the images in both views were classified as perfect (74% and 57.33% for CC and MLO views respectively). However, 3.33% of the images in MLO views were graded as inadequate (Table 6).

**Table 6.** Image quality assessment.

| Grades of image quality |  | P |  | G |  | M |  | I |  | TOTAL |  |
| --- | --- | --- | --- | --- | --- | --- | --- | --- | --- | --- | --- |
| Hospital | Views | Pts<br>NO:- | %age | Pts<br>NO:- | %age | Pts<br>NO:- | %age | Pts<br>NO:- | %age | Pts<br>NO:- | %age |
| **A** | CC | 28 | **56** | 2 | **4** | 19 | **38** | 1 | **1** | 50 | **100** |
|  | MLO | 21 | **42** | 1 | **2** | 27 | **54** | 1 | **2** | 50 | **100** |
| **B** | CC | 39 | **78** | 2 | **4** | 9 | **18** | 0 | **0** | 50 | **100** |
|  | MLO | 29 | **58** | 5 | **10** | 15 | **30** | 1 | **2** | 50 | **100** |
| **C** | CC | 44 | **88** | 2 | **4** | 3 | **6** | 1 | **2** | 50 | **100** |
|  | MLO | 36 | **72** | 1 | **2** | 10 | **20** | 3 | **6** | 50 | **100** |
| **ALL** | CC | 111 | **74** | 6 | **4** | 31 | **20.66** | 2 | **1.33** | 150 | **100** |
|  | MLO | 86 | **57.33** | 7 | **4.66** | 52 | **34.66** | 5 | **3.33** | 150 | **100** |

### Comparison of the National DRL (this study) with other established DRLs

Different countries set their DRLs for variable CBT as shown in Table 7. The DRL/woman for Malaysia was set for CBT ranges (20-39, 40-59 and 60-99mm) similar to the one in the current studies (7-39, 40-59 and 60-99mm). The respective DRL values for the two countries were 1.13, 1.52 and 1.87mGy for Malaysia and 1.57, 1.78 and 2.34mGy for the Ugandan Screening mammogram. The DRL/woman for diagnostic in this study (1.8, 2.05 and 2.57mGy) was higher than those for screening in Malaysia for a similar CBT.

**Table 7.** Comparison of the National DRLs in this study with other established DRLs.

| Source of data/country | NO:-<br>women | CBT<br>(mm) | DRL/view |  | DRL/woman |
| --- | --- | --- | --- | --- | --- |
|  |  |  | CC | MLO |  |
| **The current study (Uganda);**<br>**Screening** | **150** | **7-39** | **1.5** | **1.66** | **1.57** |
|  |  | **40-59** | **1.78** | **1.87** | **1.78** |
|  |  | **60-99** | **2.18** | **2.22** | **2.34** |
| **The current study (Uganda);**<br>**Diagnostic** | **150** | **7-39** | **1.7** | **1.91** | **1.83** |
|  |  | **40-59** | **2.0** | **2.09** | **2.05** |
|  |  | **60-99** | **2.63** | **2.81** | **2.57** |
| O'Leary et al [20], Ireland | 1010 | 40-68 | 1.45 | 1.56 | --- |
| Karsh et al[21] | 200 | 60-70 | 1.15 | 1.24 | 1.18 |
| Australia, 2017 | ---- | 30-40 | --- | -- | 1.31 |
|  |  | 40-45 | --- | -- | 1.5 |
|  |  | 45-55 | --- | -- | 1.67 |
|  |  | 55-60 | --- | -- | 2.06 |
| Mohd et al [19], Malaysia | 87 | 20-39 | --- | -- | 1.13 |
|  |  | 40-59 | -- | -- | 1.52 |
|  |  | 60-99 | -- | -- | 2.87 |
| Dzidzornu et al [22], Ghana | 979 | 3-100 | 1.6 | 2.4 | --- |
|  |  | 55-60 | 2.0 | 2.5 | --- |

## Discussion

DRL is an optimum range of doses that is safe for patients to undergo examination without losing the diagnostic value of the image and it must be established for each imaging modality to help detect the unusual level of doses given to patients [13].

This study determined and compared the NDRLs for digital mammography with those of other countries using data from 60% the total digital mammography machines in Uganda.

The digital mammography machines in 2/3 of the hospitals were Senographe and Siemens in 1/3 Hospitals in Uganda.

All hospitals in this study used a large focal spot size (0.3 mm) and tungsten/Rhodium (W/Rh) anode target/filter combination to acquire mammography images for both screening and diagnostic mammography. Modern mammography systems most frequently use a nominal focal spot size of 0.3 mm for regular mammography and 0.1 mm for magnification procedures [23].

The mean age of the patients recorded in this study for the screening and diagnostic mammography (50.28±9.32 and 47.45±13.45 years respectively) was slightly lower than the mean ages in other countries including Ghana {(54 ± 10.0 years) [24]}, {(56 ± 10.0 years) [25]} and Australia {(60 ± 7.9 years) [26]. Furthermore, the patients for screening were much younger than those for diagnostic mammography with age ranges of 15-84 years and 33-76 years respectively. The young age patients for the diagnostic mammogram were probably because of the younger age of distribution of breast cancer in Uganda. However, 15 years of age was below the recommended minimal age (25 years) of performing mammography in symptomatic patients in Uganda [27].

This study found a strong positive relationship between AGD and some exposure parameters. The relationships were comparable in both views of screening and diagnostic mammography. The findings recorded are in agreement with a report by Ko et al [28] which indicated mAs, and kVp correlated positively with CBT.

The National DRL values for diagnostic mammography obtained in this study were well higher than those for screening mammography for the corresponding CBT ranges and views. This could be due to the younger age of women turning up for diagnostic mammography in this study. The younger women have a greater proportion of breast glandular tissue than fatty tissue, which requires higher x–rays exposure, hence higher DRL values in diagnostic mammography than in screening mammography. This finding is consistent with a study by O’Leary & Rainford (Screening service DRL at 55–65 mm CBT: 1.75 mGy (screening) vs. 2.4 mGy (symptomatic) [29].

The national DRL in the current study was comparable with those in Malaysia [19]. However, most countries set their DRLs for a single narrow CBT range as in Malta which was set at a CBT range of 50-70mm, while Sudan [30], and Ireland [31] set their DRLs at 55-65mm CBT range The comparison of DRLs with those of other countries is therefore challenging due to the lack of a standard CBT range used for setting the DRLs.

The overall image quality of more than 50% was classified as perfect (74% and 57.33% for CC and MLO projections respectively) and less than 3% (1.33%) was inadequate in a CC projection. One of the hospitals had an inadequate image quality of up to 6%. While it was 6.2% according to O’Leary et al in Ireland [20]. To effectively use PGMI, a minimum of 50% of audits of 50 randomly selected cases graded as P or G categories (75% desirable), P or G or M categories (97% desirable) and the repeat rate should be less than 3% of consecutive images to be classified as “Inadequate”

## Conclusion

There was a positive relationship between the AGD and exposure factors (mAs, kVp), ESD. The patient-based NDRL values for digital screening and diagnostic mammography at different compressed breast thickness ranges in Uganda have been proposed for the first time providing valuable insights into the radiation dose status.

The established NDRLs in this study were comparable to the NDRL values for some few countries which set their NDRLs at a similar compressed breast thickness.

### Recommendation

The NDRL values in mammography should be specific to compressed breast thickness ranges, mammographic views, and indications for mammography (screening and diagnostic mammography) as significant variations in their DRL values were shown in this study.

## Data Availability

All relevant data are within the manuscript and its Supporting Information files.

## Acknowledgement

We acknowledge Dr. Nassanga Rita, Dr. Mubuuke Roy, Dr. Bugeza Samuel and Professor Dan Dell who contributed in their various capacities towards this study.

## Supporting information

**S1 appendix:** PGMI-Image quality assessment for mammography.

## Notes

### Competing Interest Statement

The authors have declared no competing interest.

### Funding Statement

The author(s) received no specific funding for this work.

### Author Declarations

Ethical approval to conduct the study was obtained from the Institutional Review Board (IRB) of the School of Medicine, Makerere University College of health sciences (protocol reference number: Mak-SOMREC-2023-562).

